# Enhancing competency in clinical trials management: Findings from a multicountry trial coordinators’ interventional training program

**DOI:** 10.64898/2026.03.03.26347517

**Authors:** Dawit Asmamaw Ejigu, Abebaw Fekadu, Eyasu Makonnen, Almari Conradie, Brenda Okech, Jennifer Lehrman, Rahel Birhane, Mahnaz Vahedi, Tsegahun Manyazewal

**Author notes:** **Address correspondence**. Dr Dawit Asmamaw Ejigu. Center for Innovative Drug Development and Therapeutic Trials for Africa (CDT-Africa), College of Health Sciences, Addis Ababa University, P.O. Box 9086, Addis Ababa, Ethiopia; Department of Pharmacology, St. Paul’s Hospital Millennium Medical College, Addis Ababa, Ethiopia. **Funding:** Gates Foundation ((Grant No.: INV-055202).

## Abstract

**Background:** Clinical research coordinators play a crucial role in ensuring the scientific rigor, regulatory compliance, and operational integrity of clinical trials. However, in Africa, they often lack access to structured, competency-based training, especially in operational, regulatory, and trial management domains. This study evaluated the effectiveness of a comprehensive training intervention designed to standardize and enhance core competencies of clinical trial coordinators.

**Methods:** We conducted a prospective pre-post interventional study among cohorts of clinical research professionals completing a 10-week, internationally-accredited, Moodle-based clinical trial operations training program aligned with the Joint Task Force Core Competency Framework, covering 10 lessons and 25 domains. Self-reported competence was evaluated at baseline and post-training. Data analyses included paired t-tests for aggregate scores, McNemar’s exact test for domain-level proportions, multivariable logistic regression for predictors of improvement, and Cohen’s *d* for effect size.

**Results:** Among the 166 participants enrolled from 19 African countries and completed the pre-training survey, 152 who completed the program and post-training survey were included. The training significantly increased the mean aggregate competence from 12.24±7.85 (out of a maximum of 25) to 23.35±2.73 (mean difference: 11.11; 95% CI 9.86-12.36; *p<0*.*001*; Cohen’s *d*=1.41). Score variance decreased, with the median score increasing from 12.0 (IQR: 6.0-19.0) to 24.5 (IQR: 23.0-25.0). All 25 domains improved (p<0.001), with the largest gains in complex, low-baseline domains: managing external partners (+59.2%), project management (+58.6%), financial management (+55.3%), and trial close-out (+57.2%). (+57.2%). Ethical principles and informed consent that had high baseline competence reached near-universal levels at 99.3% and 98.7%, respectively. No differences were observed by country or gender (*p>0*.*05*).

**Conclusion:** Structured, competency-based training strengthens clinical trial coordinators’ capabilities, particularly in technical and administrative domains that are often overlooked. Accredited, framework-aligned clinical trial training programs promote consistent trial quality, strengthen research capacity, and sustain excellence in clinical trial delivery.

**WHAT IS ALREADY KNOWN ON THIS TOPIC:**

- Clinical research coordinators play a crucial role in ensuring the scientific rigor, regulatory compliance, and operational integrity of clinical trials

**WHAT THIS STUDY ADDS:**

- The study evaluated the effectiveness of a comprehensive training intervention designed to standardize and enhance core competencies of clinical trial coordinators in Africa, where they often lack access to structured, competency-based training

**HOW THIS STUDY MIGHT AFFECT RESEARCH, PRACTICE OR POLICY:**

- This study should encourage the design and delivery of internationally-accredited, Moodle-based clinical trial operations training programs in Africa that enhance clinical trial competency.

## Background

The complexity, diversity, and safety requirements of clinical trials have rendered their implementation more challenging and dependent on the operational and regulatory competencies of the clinical trial workforce [1,2]. While structured education and competency frameworks have been well established in developed countries to standardize clinical trials training and the role of the workforce [3]. These frameworks have yet to be implemented sufficiently in developing countries, including those in Africa [4].

Clinical trial activity in Africa has surged rapidly in recent years, driven by increasing disease burden, research infrastructure, and global investment in clinical trials [5,6], but not always matched by systematic capacity-building for the essential trial workforce. Studies that evaluated clinical trial capacity in Africa have identified gaps in training and practical experience among trial researchers, especially in trial management and coordination [7,8]. These gaps may lead to delays, protocol deviations, and regulatory non-compliance, which potentially discourage conducting trials in Africa.

Structured learning has been shown to enhance knowledge, confidence, and performance of the trial workforce [9]. Recent evaluations of clinical research training programs, including competency-based approaches and formal competency-based curricula in clinical trials, have been associated with improvements in trial operations, increased trust in trial sponsors, and improved regulatory understanding and coordination skills among trial researchers in various fields [10]. However, there remains no systematic, multidisciplinary, outcome-based evaluation of clinical trial workforce training in Africa.

In response to these gaps, a novel 10-week competency-based clinical trial operations training program using an online platform was developed and delivered for clinical trial coordinators working in different African clinical research environments [11]. The program covers core competencies in designing, conducting, managing, and reporting clinical trials, areas identified as weaknesses in previous assessments of clinical research workforce capability.

The aim of this study was, therefore, to assess the effectiveness of the clinical trial operations training program in improving clinical trial competency of study coordinators in Africa.

## Methods

### Design and program

This was a single-group, pre-post evaluation of the competency-based clinical trial operations training program, a multi-country initiative for training clinical trial coordinators in Africa. The study involved a cross-sectional analysis of baseline survey data from participants enrolled in the training, followed by a paired pre- and post-training observational assessment to determine changes in trial coordination knowledge. Each participant served as their own control, enabling direct comparison of competencies before and after completing the program.

The design and implementation strategy of the training program is described elsewhere [11]. In brief, the training is a 10-week competency-based course for African study coordinators and other researchers. In the learning process, the program uses cloud-based platforms, such as VoiceThread for interactive, offline-accessible content, Moodle for distribution of resources, Zoom for live tutorials and mentoring, and competency assessment, including quizzes, discussion forums, tutorials, and group assignments. Guided by pre- and post-intervention teaching design, the course integrates cognitive, constructivist, and humanistic teaching theories and emphasizes problem-based learning, mentoring, and social engagement. Participants spent 8-12 hours a week on ten weekly lessons covering Introduction to Clinical Trials Operations, Study Design and Protocol Development, Project and Financial Management, Conducting a Trial (Part 1), Conducting a Trial (Part 2), Closing out and Reporting a Trial, Working with External Partners, Quality Systems, Audits & Inspections, and Pharmacovigilance. Participants’ competencies are evaluated using quizzes, discussion forums, tutorials, and group assignments, with certification awarded upon achieving a minimum cumulative score of 70%. The program was developed in collaboration with academic and product development partners including the Faculty of Capacity Development (FCD), Foundation for Innovative New Diagnostics (FIND), Medicines for Malaria Venture (MMV), International AIDS Vaccine Initiative (IAVI), Program for Appropriate Technology in Health (PATH), and the Special Programme for Research and Training in Tropical Diseases (TDR), among others. The program was initially developed by FCD and the global partners and was then successfully transferred to the Center for Innovative Drug Development and Therapeutic Trials for Africa (CDT-Africa), Addis Ababa University, Ethiopia, for regional implementation. Following a rigorous evaluation process, the program received international accreditation from a European-based international accreditation agency.

### Participants

Participants were study coordinators recruited for the clinical trial operations training program. Eligibility required enrolment in the program and completion of pre- and post-course competency assessments. The program targeted African study coordinators or individuals aspiring to work in this field, with continent-wide merit-based selection prioritizing applicants with prior experience as a study coordinator, investigator, co-investigator, study manager, or assistant, and preferably with academic qualifications in medicine, nursing, pharmacy, biomedical sciences, or related fields.

### Data collection

Participants completed a validated 25-item competency assessment before and after the training, along with a baseline comprehensive set of open and closed-ended questions. The cross-sectional baseline survey collected demographic and professional characteristics, previous training and experience in study coordination, clinical trial performance, quality system implementation, site operational capacity, scientific dissemination, and managing challenges at sites.

As part of the pre- and post-training competency assessment, participants completed a closed-ended questionnaire comprising 25 questions, which assessed their self-confidence in the ten areas of learning, as described above, to relate their perceived sense of competence. The competency assessment questionnaire was structured to generate a score for each of the 25 competency items, assigning 1 for “competent” and 0 for “not competent”. Given the high competency standards required in the clinical trials landscape, participants rated each question on a 5-level scale: self-reported competency ratings of “excellent” or “good” were classified as competent (1), whereas ratings of “fair,” “some,” or “poor” were classified as not competent (0).

### Outcome measures and variables

The primary outcome measure was the aggregate post-training competency level of participants in coordinating clinical trials, defined as the composite competency score achieved post-training (0-25) as compared to pre-training, analyzed using a paired t-test and confirmed using the Wilcoxon signed-rank test.

Secondary outcomes included: 1) domain-by-domain change in proportion of competency, defined as the improvement score obtained in McNemar’s test. 2) High competency proficiency performance, defined as the attainment of the highest competencies post-training, calculated by subtracting the pre-training competence score from the post-training competence score in a multivariable linear regression. 3) Association between competency improvement and characteristics of study participants, where the independent predictor variables included ten general characteristics of the participants: 1) Experience as a clinical trial coordinator, 2) Prior training in clinical trials, 3) Experience in clinical trials, 4) Clinical trial employment status, 5) Experience in scientific presentation, 6) Gender, 7) Country, 8) Quality system present at site, 9) Risk management system present at site, and 10) Baseline competency score.

### Statistical analysis

Descriptive statistics were used to summarize the characteristics of the study participants in the analysis of their baseline characteristics. The categorical variables were presented as frequencies and percentages. The continuous variables were summarized using mean values with standard deviations or mean values with interquartile ranges.

Pre- and post-test differences were assessed using parametric (paired t-tests), and the magnitude of the effect was estimated by Cohen’s d. Competency was assessed as a binary outcome (competent vs. not competent) and as composite competency scores ranging from 0 to 25. For each of the 25 competencies, paired odds ratios were compared using the McNemar test, and the improvement in competency was summarized by pre-competency (%), post-competency (%), absolute improvement (%), and p-value of the McNemar test. The overall improvement in competency was assessed on the basis of the total composite scores of the participants, which reported the mean pre-score, mean post-score, mean improvement, and effect size. Statistical significance was defined by *p<0*.*05*.

In order to identify predictors of the outcomes of training, subgroup analyses examined changes based on level of experience and previous training. Multivariable logistic regression models were developed to assess participant characteristics associated with post-training competence and competence improvement. All variables were fed into the models at the same time, and the regression coefficients, probability ratios and 95-percent confidence limits were reported. Multiple linear regression was also used to identify predictors of overall score improvements.

### Ethical considerations

The Scientific and Ethics Review Committee of the Center for Innovative Drug Development and Therapeutic Trials for Africa, Addis Ababa University, approved the study. Informed consent was obtained from study participants. The study adhered to the Declaration of Helsinki.

## Results

### Participant characteristics at baseline

A total of 166 participants from 19 African countries, comprising equal (50%) representation of females and males, were enrolled at baseline. Most of the participants were from Kenya (n = 36; 21.7%), Ethiopia (n = 28; 16.9%), and Uganda (n = 25; 15.1%. The participants were drawn from two cohorts: namely, 2023 (n = 91, 54.8%) and 2025 (n = 75, 45.2%) cohorts. At baseline, 89 participants (53.6%) reported previous experience as site coordinators with a median duration of 24 months (range: 1-96). Of these participants, 74 (44.6%) reported using operational performance indicators, 25 (15.1%) had designed a clinical trial quality system, and 86 (51.8%) reported the presence of formal risk management documentation at their sites (**Table 1**).

**Table 1:** Baseline characteristics of study participants.

|  | Characteristics | Number | % |
| --- | --- | --- | --- |
| Distribution by sex | Sex |  |  |
|  | Female | 83 | 50 |
|  | Male | 83 | 50 |
| Distribution by country | Country |  |  |
|  | Kenya | 36 | 21.7 |
|  | Ethiopia | 28 | 16.9 |
|  | Uganda | 25 | 15.1 |
|  | Nigeria | 17 | 10.2 |
|  | Tanzania | 13 | 07.8 |
|  | South Africa | 09 | 5.4 |
|  | Zambia | 06 | 3.6 |
| | Others ($\leq 5$, n = 12) * | 32 | 19.3 |
| Ever worked as research coordinator | Coordinator experience |  |  |
|  | Yes | 89 | 53.6 |
|  | No | 77 | 46.4 |
| Prior clinical trial coordination course | Prior course |  |  |
|  | Yes | 17 | 10.2 |
|  | No | 149 | 89.8 |
| Ever designed clinical trial quality system | Designed quality system |  |  |
|  | Yes | 25 | 15.1 |
|  | No | 141 | 84.9 |
| Use of operational indicators at site | Use indicators |  |  |
|  | Yes | 74 | 44.6 |
|  | No | 92 | 55.4 |
| Risk management documentation at site | Risc documentation |  |  |
|  | Yes | 86 | 51.8 |
|  | No | 80 | 49.2 |
| NB: *Burkina Faso (5), Rwanda (4), Democratic Republic of Congo (4), Liberia (4). Sierra Leone (3), Senegal (2), Malawi (2), Mozambique (2), Botswana (2), Ghana (2), Gabon (1), Zimbabwe (1) |  |  |  |

### Aggregate pre-post-training performance

Of the 166 participants who initially enrolled and completed the pre-course survey, 154 completed the training successfully, and 152 replied to the post-course survey. The pre-post analysis was, therefore, based on the data of 152 participants, 91.6% of the initially enrolled participants.

A statistical analysis of the aggregate assessment of participants’ pre- and post-training competence underscores significant improvements in self-reported clinical trial coordination competencies following the training intervention.

At baseline, the participants demonstrated a moderate and highly variable range of self-reported basic competencies, with a mean score of 12.24 ± 7.85 out of a maximum of The baseline median score was 12.0 (IQR: 6.0 −19.0) and a full range of 0-25. After completing the course, there was a significant shift in the participants’ aggregate competencies. The mean total score increased significantly, with a post-training mean course score of 23.35 ± 2.73, which shows a significantly higher mean increase of 11.11 points (95 CI 9.86-12.36, *p<0*.*001*). The effect size of the intervention was significantly large, with Cohen’s *d* of 1.41. In addition to the mean increase, the variance was also significantly reduced, with the median score increasing from 12.0 (IQR: 6.0-19.0) before training to 24.5 (IQR: 23.0-25.0) after training.

### Domain-specific pre-post-training performance

The baseline (pre-training) data revealed specific areas of relative weakness, in particular financial management (19.7%), protocol development (24.3%), and external partner governance (31.6%). On the other hand, ethical principles and informed consent were the areas of highest initial competence (73.7% and 78.9%, respectively).

An item-by-item analysis using McNemar’s exact test confirmed that the improvements were statistically significant in every single domain evaluated (p<0.001 for all items) (**Table 2**). The most significant increases in competencies were found in complex project management tasks, such as managing external partners (+59.2%), project management of trial sites (+58.6%), and coordinating trial closeout (+57.2%). Even in domains where initial competence was high, such as informed consent, significant further improvements were made, with almost universal competence (98.7%) achieved after the course.

**Table 2:**
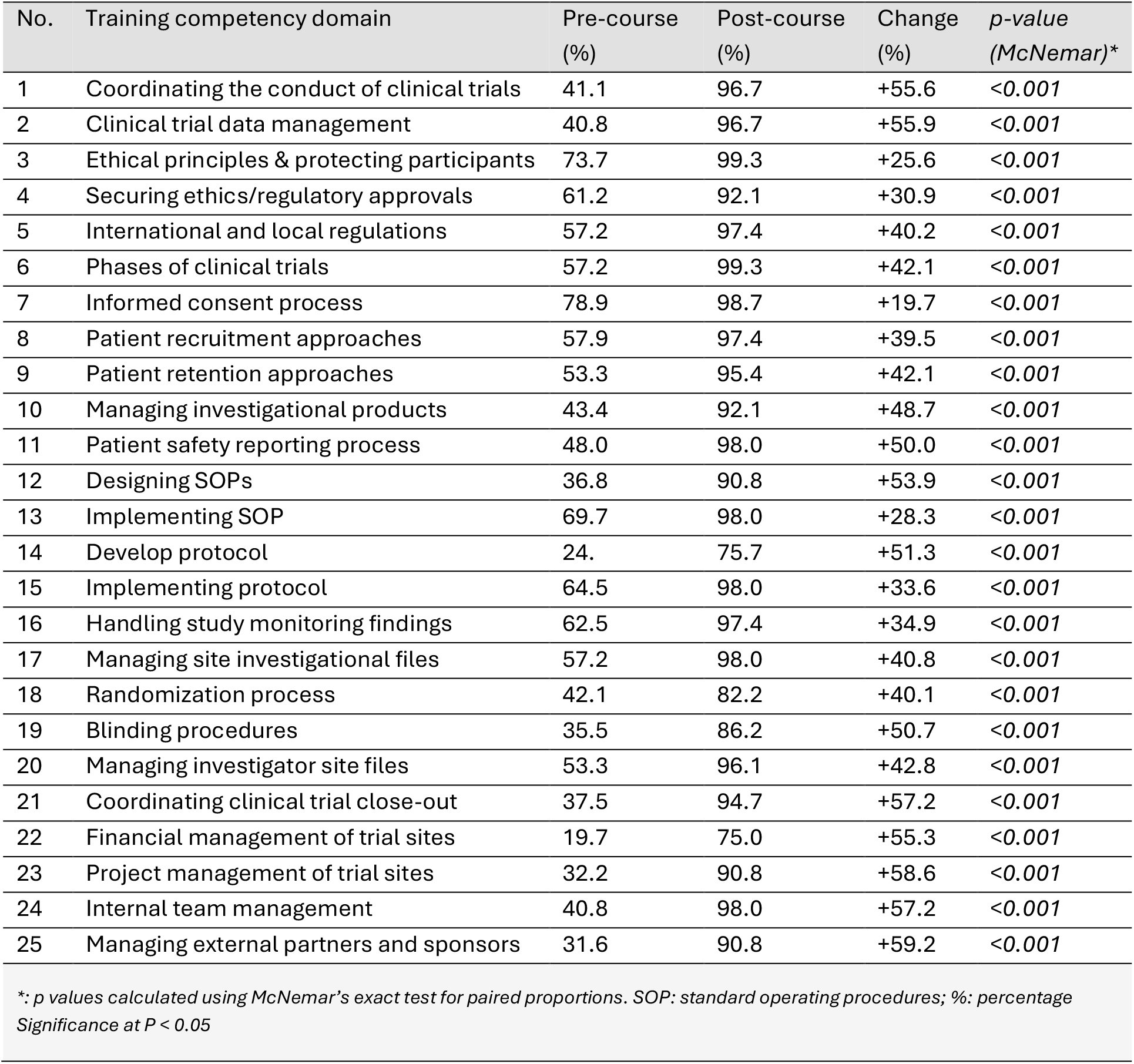
Competence proportions and statistical comparisons.

### Factors associated with competency improvement

In the multiple linear regression and subgroup comparisons, which primarily represent participants’ demographic background, prior experience, and course satisfaction, gender and country did not significantly affect baseline scores or the rate of improvement post-training (p>0.05), which demonstrates that the training intervention was equally effective across genders and countries (**Table 4**).

**Table 4:** Multiple regression analysis of the factors associated with the change in competency scores.

| Variable | Predictor level | Coefficient ($\beta$) | Std. Error | t-value | p-value |
| --- | --- | --- | --- | --- | --- |
| Intercept | - | 16.13 | 1.29 | 12.55 | **<0.001** |
| Worked as a coordinator | Experience | -3.61 | 1.24 | -2.92 | 0.004 |
| Past coordination training | Prior training | -2.98 | 2.02 | -1.47 | **0.143** |
| Maintained quality systems | Technical experience | -3.65 | 1.34 | -2.72 | **0.007** |
| Designed quality systems | Advanced experience | -1.92 | 1.70 | -1.13 | 0.260 |
| Sex (female) | Gender | 0.15 | 1.16 | 0.12 | 0.906 |
| Key performance indicators use | Site practice | -0.40 | 1.21 | -0.33 | 0.739 |
| Significance at $P < 0.05$ | | | | | |

In terms of the impact of experience on training outcomes, while the overall training effect was very strong, regression analysis shows that individual learning pathways are significantly influenced by previous work experience. In particular, previous work as a clinical coordinator and prior maintenance of quality systems were the most reliable predictors of baseline knowledge and subsequent improvements. Regarding baseline competency and prior experience, participants who had prior experience working as clinical trial coordinators or had experience in maintaining quality systems had significantly higher baseline competence scores when they started the training (*p = 0*.*009* and *p = 0*.*003*, respectively). At baseline, on average, experienced coordinators scored 5.38 points higher than those without such experience (mean score: 14.64 vs. 9.26) (**Table 4**).

In terms of the magnitude of improvement, linear regression analysis computed using the change score (post-training minus pre-training) as the dependent variable demonstrated that prior experience was significantly negatively associated with the magnitude of improvement (*p< 0*.*01*). Participants with previous coordinator experience showed a mean improvement of 8.63 points, whereas those without prior experience improved by 14.8 points (**Table 5**). This pattern suggests a ceiling effect, which indicates that the training yielded greater gains among novice coordinators and substantially narrowed the competency gap between entry-level and experienced participants.

**Table 5:** Comparative outcomes by professional experience, triangulating the paired t-test results with prior coordination experience.

| Experience group | N | Pre-training mean, SD | Post-training mean, SD | Improvement mean | *P-value* |
| --- | --- | --- | --- | --- | --- |
| No prior experience | 68 | 9.26 ± 7.39 | 23.44 ± 2.49 | +14.18 | <0.001 |
| Prior experience | 84 | 14.65 ± 7.41 | 23.27 ± 2.93 | +8.63 | <0.001 |
| Total in cohorts | 152 | 12.24 | 23.35 | + 11.11 | <0.001 |
| *Significance at $p < 0.05$* |  |  |  |  |  |

## Discussion

A structured, high-quality, virtual training program for clinical trial coordinators was evaluated for its impact on improving self-reported competency, which showed a significant improvement after completion of the training. The very large magnitude of the effect and the general increase in proficiency across all 25 competency areas suggest that targeted training may effectively bridge the knowledge gap for clinical researchers. These findings are in line with recent international literature, which highlights the shift towards competence-based training as a means of ensuring the quality of trials and the safety of participants [12].

A major finding is the difference in baseline competencies. More than 70% of the participants were initially most confident in ethical principles and informed consent, which might be attributed to the mandatory nature of good clinical practice certification in most research environments. However, the baseline scores were particularly weak in specialized administrative and technical areas such as financial management and external partner management. This is consistent with recent studies and shows that, while coordinators often have a robust clinical background, they often lack the entrepreneurial and project management skills needed for the complex landscape of modern clinical trials [13, 14].

The most significant improvements, often exceeding by 50%, occurred in areas related to the clinical trial site operations, including internal team management, data management, and trial closure. The significant change shows that the training has effectively addressed these knowledge and operational gaps, and recent evidence from multicenter studies suggests that such comprehensive training is essential to reduce protocol violations and improve data integrity, especially in resource-limited environments where formal study coordinator training programs are limited [15,16].

In addition, the significant reduction in the variance of scores after the training intervention, marked by reduced interquartile range, underlines the course-leveling effect of the program. By bringing most participants to a stronger level of competency, the program has proven its usefulness in a wide range of cohorts with different levels of experience. This consistency is vital to standardizing the implementation of clinical trials in the regional and global research networks [17,18].

The findings show how inclusive the training program is. The training successfully raised both experienced and inexperienced participants to nearly equal high levels of post-course competence, even though the overall “very large” Cohen’s *d* seen in the aggregate analysis was primarily driven by significant gains among novice coordinators. The program’s ability to standardize knowledge and skills across a diverse professional group is reflected in this leveling effect. Additionally, baseline competency scores and the degree of post-training improvement were not significantly impacted by gender or geographic location, suggesting that the intervention was equally effective for both sexes and countries.

It must, however, be recognized that self-reported competence measures reflect self-reported efficiency. Self-efficacy is a strong predictor of professional performance and retention, but it is not always a direct proxy for objective competence. Recent trends in clinical research training favor adding longitudinal performance metrics and work-based assessments to these evaluations to ensure that the improvements reported here are translated into long-term operational excellence [19–22].

## Conclusion

In this study, structured, competency-based training substantially enhances the capabilities of clinical trial coordinators, demonstrating a significant improvement after completion of the training, particularly in technical and administrative domains that are often overlooked in traditional clinical training. These findings support the potential value of accredited, framework-aligned training programs in promoting consistent trial quality across diverse research settings. Formalized training for clinical trial coordinators may represent an important component of strategies aimed at strengthening research capacity and sustaining excellence in clinical research delivery.

## Data Availability

All data produced in the present study are available upon reasonable request to the authors

## Declarations

### Data availability

The datasets used and/or analyzed during the current study are available from the corresponding author upon reasonable request.

## Acknowledgments

The authors thank the Center for Innovative Drug Development and Therapeutic Trials for Africa (CDT-Africa), Addis Ababa University, for successfully hosting and managing the project.

## Funding statement

This effort was made possible through funding from the Gates Foundation (Grant No.: INV-055202). The content is solely the responsibility of the authors.

## Authors’ contributions

Study design and conception: DAE, TM, AF, EM, AC, BO, JL, MV, Funding acquisition: DAE, AF, EM. Study implementation and data analysis: DAE, TM, AF, EM, RB. Draft the Manuscript: DAE, TM. Reviewed and revised the manuscript: DAE, TM, AF, EM, AC, BO, JL, RB, MV, All authors read and approved the final manuscript for publication.

## Competing interests

All authors report no conflict of interest.

## Notes

### Competing Interest Statement

The authors have declared no competing interest.

### Author Declarations

The Scientific and Ethics Review Committee of the Center for Innovative Drug Development and Therapeutic Trials for Africa, Addis Ababa University, approved the study

